# Hereditary Hemorrhagic Telangiectasia in Uruguay: Epidemiologic and clinical features of the evaluated population

**DOI:** 10.1101/2025.08.18.25333772

**Authors:** Z. Criscuolo, C. Chiesa, G. Losada, B. Marsiglia, L. Matta, R. Nogara, H. Pereira, S. Rodriguez, R. Mezzano, S. Ruiz

**Author notes:** Corresponding author: Dr. Santiago Ruiz, Laboratory of Metabolic diseases and Aging, Institut Pasteur de Montevideo, Montevideo, CP 1400, Uruguay.

## Abstract

**Background:** Hereditary Hemorrhagic Telangiectasia (HHT) is a rare autosomal dominant vascular dysplasia, characterized by mucocutaneous telangiectasias and visceral arteriovenous malformations (AVMs). Despite an estimated global prevalence of 1 in 5,000, HHT remains underdiagnosed in many regions. Prior to this study, no epidemiologic data were available for Uruguay.

**Objective:** To describe the epidemiologic and clinical characteristics of HHT patients in Uruguay, estimate the national prevalence of the disease, assess adherence to international screening guidelines, and diagnostic delays.

**Methods:** A cross-sectional observational study was conducted in Uruguay using data from the national HHT reference registry. Patients who met the Curaçao criteria or had a confirmed pathogenic genetic variant were included. Data were obtained through a standardized telephone survey and medical record review. Variables included demographic data, clinical manifestations, diagnostic workup, and treatment. Descriptive and bivariate analyses were performed using SPSS software.

**Results:** The registry included 134 patients, of whom 90 were surveyed. Estimated HHT prevalence in Uruguay was 3.83 per 100,000 inhabitants (95% CI, 3.26 - 4.61). The mean age was 48.2 years (SD ± 18.3), with a female-to-male ratio of 1.73:1. Epistaxis affected 88.9% of adults, with more than 50% classified as moderate-to-severe and had anemia. Common treatments included oral (60.7%) and intravenous (41.6%) iron, tranexamic acid (24.7%), and propranolol (11.2%). Pulmonary, cerebral, hepatic and digestive arteriovenous malformations were present in 20%, 15%,19% and 50% of patients, respectively. The mean diagnostic delay was 5.7 years (SD ± 10.6). Only 22% of patients completed the recommended screening.

**Conclusion:** HHT remains underdiagnosed and undertreated in Uruguay, with significant diagnostic delays and low adherence to international screening guidelines. This first national epidemiological study underscores the urgent need for a formally recognized national referral center to ensure high-quality, multidisciplinary care and to raise disease awareness, with the aim of reducing preventable complications.

## Background

Hereditary Hemorrhagic Telangiectasia (HHT), also known as Rendu-Osler-Weber syndrome, is an autosomal dominant vascular dysplasia with a prevalence of 1 in 5,000 to 10,000 individuals worldwide (1). Its main clinical manifestations include recurrent and spontaneous epistaxis (95%), mucocutaneous telangiectasias (90%) located in the face, mouth (lips and tongue), hands, gastrointestinal tract (80%), and arteriovenous malformations (AVMs) in organs such as liver (74%), lungs (50%), and brain (25%) (2,3). Other relevant manifestations that also affect the quality of life are related to blood loss (4). Epistaxis and bleeding in the digestive tract can cause iron-deficiency anemia (5) that affects 50% of the patients and it is associated with high morbidity and mortality (6). Some of the most important complications of the AVMs are stroke and brain abscess due to lung MAVs, cerebral hemorrhage and seizures due to brain AVMs and cardiac failure or portal hypertension due to liver AVMs (7). The phenotype and complications of the disease tend to worsen along patients’ age and although some reports state no gender propensity among HHT patients (8), others described several gender differences and severity (9).

HHT diagnosis is based on the presence of at least three of the four Curaçao criteria: spontaneous and recurrent epistaxis, multiple mucocutaneous telangiectasias, presence of systemic AVMs in the liver, lungs, brain and gastrointestinal tract, and a first-degree family history of HHT. Although these criteria can establish the diagnosis in most cases, disease penetrance increases with age; therefore, genetic testing is also used as a diagnostic tool, particularly in pediatric patients and in cases of uncertain diagnosis (10).

Several mutated genes involved in the transforming growth factor-beta (TGF-β) signaling pathway have been identified, specifically affecting the BMP9/10-ENG-ALK1-SMADs pathway in endothelial cells (11). HHT is genetically classified mainly in subtypes HHT1 (OMIM#187300) and HHT2 (OMIM#600376), with mutations in the genes endoglin and ACVRL1 respectively (12). Mutations in the gene SMAD4 have been described in a syndrome that combines juvenile polyposis and HHT (JP/HHT;OMIM#175050) (13,14), and mutations in the gene GDF2 in a HHT-like syndrome (subtype HHT5, OMIM#615506) (15). Additionally, two loci have been identified by linkage analyses on chromosomes 5 (HHT3) and 7 (HHT4), but their coding genes have not been identified (16,17).

A lack of knowledge of presumptive patients and awareness of HHT’s clinical features among healthcare providers can result in delayed diagnoses and increased morbidity and mortality. For instance, a delay of 25 years has been identified between the onset of the disease and the definitive diagnosis (18). Therefore, outreaching and raising awareness to reduce time and improve accuracy in the screenings and diagnostics is essential to prevent complications. Epidemiological studies are key to understanding the scope and impact of HHT.

Although there is a dedicated team actively promoting the care of HHT patients -referred to as the HHT Uruguay Reference Group and recognized by Cure HHT-a formally designated referral center endorsed by national health authorities has not yet been established to ensure and guarantee high-quality care for this population.

While a growing body of scientific literature has emerged from developed countries, there remains a marked gap in epidemiological data on HHT in Latin America, where studies are scarce or only beginning to emerge. Given the absence of national data and the underrepresentation of the region in the global literature, understanding the local epidemiological profile is essential for guiding public health policies and improving care delivery. This study aims to describe the clinical characteristics and epidemiological profile of patients with HHT in Uruguay.

## Method

### Design, population and setting of the study

A descriptive observational cross-sectional study was conducted on patients with HHT registered by the Uruguayan HHT reference group. The study was conducted in the Medical 1 Academic Unit of the Maciel Hospital in Montevideo, Uruguay. For case inclusion, individuals were required to have a clinical diagnosis of HHT. Diagnosis was based on Curaçao criteria (10). Case detection was based on the institutional registry, with data extracted and corroborated from REDCap medical records. A questionnaire was submitted to the HHT patients referring to our HHT reference group from 2020 to 2024. All patients in the registry who met the inclusion criteria were consecutively included, without any selection criteria. A consecutive sampling method was used, including all patients who were part of the HHT reference group registry and met the inclusion criteria. Prior to the collection of the data through the questionnaire, oral informed consent was obtained for the patients. For patients under 18 years old who participated in the study (n=7), progressive autonomy was considered, with prior approval from the responsible guardian.

### Questionnaire and variables considered for the study

The questionnaire dealt with demographic variables including age, sex, family history and location (capital of the country, Montevideo or interior). Clinical variables included age of onset and type of first manifestations, epistaxis and its characteristics according to the epistaxis severity score (ESS), lowest and current hemoglobin levels, presence of mucocutaneous telangiectasias, presence of pulmonary, cerebral, hepatic AVMs, and gastrointestinal telangiectasias. Then, questions concerning medical referral for HHT-related manifestations including bubble echocardiogram, chest angiography, cranial MR angiography, endoscopic studies, and hepatic doppler ultrasound. Lastly, pharmacological therapeutic questions regarding treatments, medication and interventions.

### Statistical Analysis

To estimate the prevalence of the disease, the total number of HHT-patients identified in the registry was divided into the total population of Uruguay according to the most recent national census (2023). For continuous variables, central tendency and dispersion measures were evaluated according to their distribution. The normality of the data was checked using the Shapiro-Wilk test. Normally distributed data were presented with mean and standard deviation (SD), while non-normally distributed data were presented with median and interquartile range (IQR). New variables were created for stratification into categories based on the ESS. A new variable was also created to analyze the difference between the age at the first consultation for the disease and the age at diagnosis, with the goal of obtaining an average diagnostic delay, along with its standard deviation. Data was stored in a database, processed and analyzed using IBM Statistical Package for the Social Sciences (SPSS) Statistics version 29.0.1.0.

### Ethical Aspects of the Research

Obtained data and information was only used for research purposes. The study was approved by the Ethics Committee of Maciel Hospital, Montevideo, Uruguay (code #13, 2023).

## Results

### Study population and prevalence

At the beginning of the study, the registry of the HHT reference group included 100 patients. Increased awareness generated during the telephone survey prompted additional referrals by family members, resulting in an expansion of the registry to 134 patients. Based on these data and the most recent national census conducted in 2023, the estimated prevalence of HHT in Uruguay was 3.83 per 100,000 inhabitants (95% CI, 3.26 - 4.61).

### Survey participation and baseline characteristics

A total of 89 patients were surveyed. The remaining patients could not be contacted or were added to the registry during the study period and had not yet completed data collection. Baseline characteristics are detailed in Table 1. There was a predominance of female participants (female-to-male ratio, 1.73:1), and the majority resided outside the capital, Montevideo. A family history of HHT was reported by 94.4% of participants. Two cases involved *de novo* pathogenic variants, while the remaining individuals without a family history had not undergone genetic testing.

**Table 1.** Demographic characteristics of the studied HHT population (n = 89)

| **Variable** | **Mean (SD) or Frequency (n)** |
| --- | --- |
| Age | 48.2 (18.3) |
| Age at first consultation | 29.2 (16.2) |
| Age at diagnosis | 32.9 (17.1) |
| Biological sex |  |
| Female | 63.3% (57) |
| Male | 36.7% (33) |
| Location |  |
| Interior | 54.5% (49) |
| Montevideo | 45.6% (41) |
| Family history of HHT |  |
| Positive | 94.4% (85) |
| Negative | 5.6% (5) |

The mean age of the study population was 48.2 years (SD ± 18.3). The mean age at first medical consultation related to HHT was 29.2 years (SD ± 16.2), and the mean age at diagnosis was 32.9 years (SD ± 17.1). The average diagnostic delay was 5.7 years (SD ± 10.6).

### Epistaxis severity and treatment

Epistaxis was reported in 88.9% of patients, with a mean age of onset of 17.6 years (SD ± 11.9). Although age-stratified prevalence showed a progressive increase, 61.3% of individuals had already experienced epistaxis before the age of 20, suggesting that onset beyond this age was less frequent (Table 2). More than half of the surveyed patients were classified as having moderate (n: 23, 26%) or severe (n: 21, 23%) epistaxis based on the ESS. Among patients with severe epistaxis, 90.5% had received oral iron therapy, 81% had received intravenous iron, and 52.4% had been treated with tranexamic acid (Table 2). The median lowest hemoglobin (Hb) level across the cohort was 7 g/dL (SD ± 3.7), whereas the median most recent Hb level was 12 g/dL (SD ± 2.3) (Table 3). Stratification by ESS severity revealed the following median lowest Hb levels: 12 g/dL in mild ESS, 6.7 g/dL in moderate ESS, and 5.5 g/dL in severe ESS. The corresponding most recent Hb levels were 13 g/dL, 10.5 g/dL, and 9 g/dL, respectively.

**Table 2.**
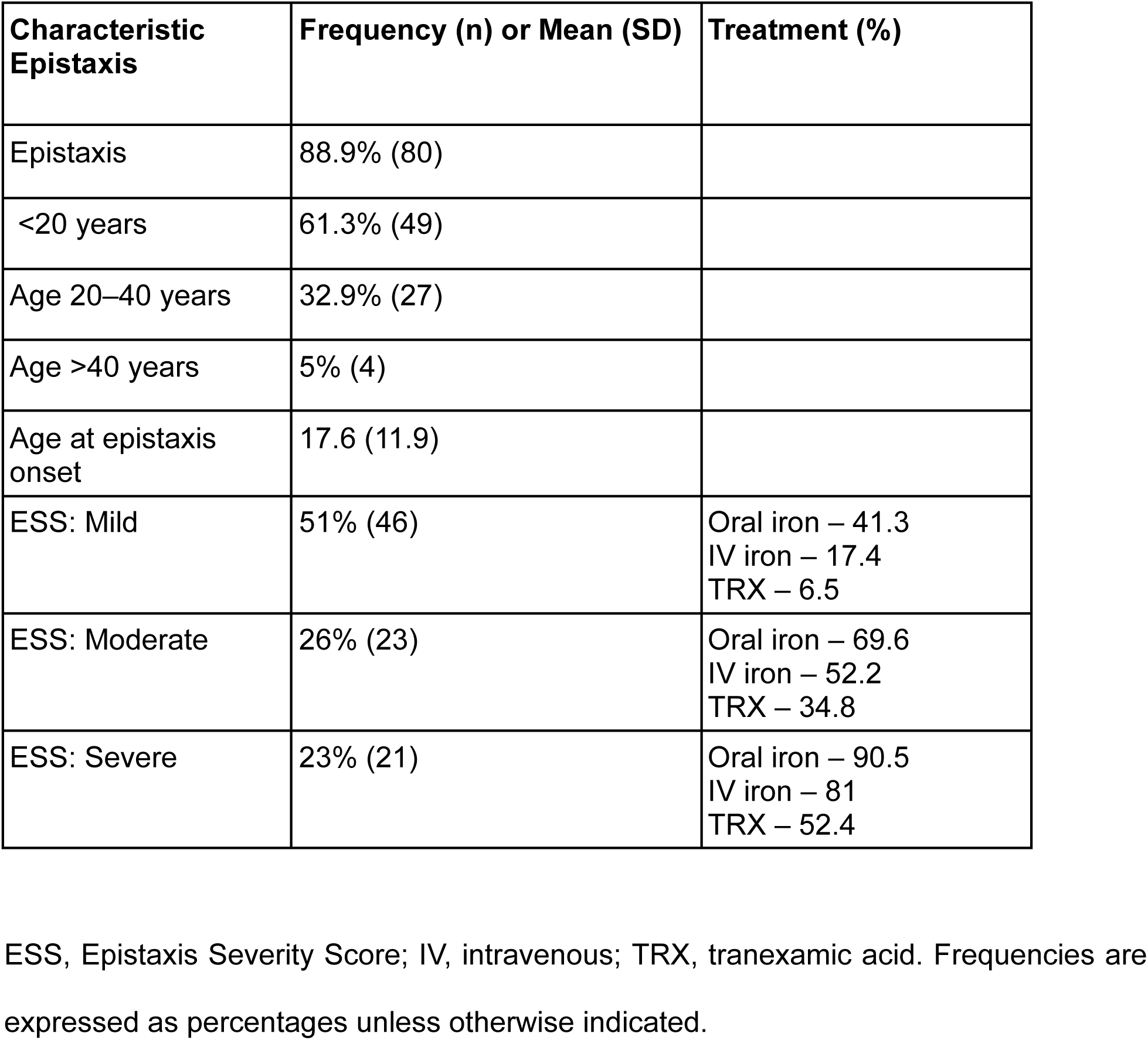
Epistaxis in the Studied Population.

**Table 3.** Most recent and lowest hemoglobin (Hb) levels in the studied population.

| **Hemoglobin (Hb)** | **Median (RI)** |
| --- | --- |
| Lowest level | 7 (5-12) |
| Most recent level | 12 (11-14) |
| ESS: Mild |  |
| Lowest Hb | 12 (6-13) |
| Most recent Hb | 13 (12-13.85) |
| ESS: Moderate |  |
| Lowest Hb | 6.7 (6-8) |
| Most recent Hb | 10.5 (10-13) |
| ESS: Severe |  |
| Lowest Hb | 5.5 (4.15-7.25) |
| Most recent Hb | 9 (7.9-12.2) |

Hb, hemoglobin; RI, reference interval.

### Clinical manifestations and organ involvement

Characteristic mucocutaneous telangiectasias (nose, lips, tongue, fingers) were observed in 90% of patients (Table 4). Visceral involvement included pulmonary AVMs in 20% of patients, cerebral AVMs in 15.7%, and hepatic AVMs in 18.9%. Telangiectasias in the upper and lower gastrointestinal tracts were identified in 34.4% and 15.6% of patients, respectively. Among patients with hemoglobin levels <10 g/dL and evidence of gastrointestinal involvement, telangiectasias were found in the upper tract in 62.5% of cases and in the lower tract in 18.75%.

**Table 4.** Clinical manifestations in the population.

| Clinical manifestation | Frequency (%) |
| --- | --- |
| Skin and mucus telangiectasias | 90 |
| Pulmonary AVMs | 20 |
| Cerebral AVMs | 15.7 |
| Hepatic AVMs | 18.9 |
| Upper digestive tract telangiectasias | 34.4 |
| Lower digestive tract telangiectasias | 15.6 |

Complications related to pulmonary AVMs included transient ischemic attack (11.1%), ischemic stroke (12.2%), and cerebral abscess (2.2%), totaling 23 reported events (Table 5).

**Table 5.** Pulmonary AVM complications in the studied population.

| **Pulmonary AVM complication** | **Frequency (%)** |
| --- | --- |
| Stroke (CVA) | 12.2 |
| Transient ischemic attack (TIA) | 11.1 |
| Brain abscesses | 2.2 |

### Screening and Diagnostic Evaluation

Complete screening, defined as having undergone bubble contrast echocardiography, cerebral angio-MRI or angio-CT, and hepatic Doppler ultrasound, was performed in 21,1% of patients (n = 19). An additional 8,9% (n = 8) underwent partial screening that included both bubble echocardiography and cerebral AVM evaluation.

The most commonly performed studies were cerebral angio-MRI or angio-CT (53.3%), thoracic angio-CT (41.6%) and bubble echocardiography (41.1%). Gastrointestinal evaluation included upper endoscopy (41.6%) and colonoscopy (42.7%). Hepatic Doppler ultrasound was performed in 38.1% of patients (Table 6).

**Table 6.** Screening tests performed.

| Study performed | Frequency (% , n) |
| --- | --- |
| Bubble echocardiogram | 41.1 (37) |
| Chest CT angiography | 41.6 (37) |
| Brain MRI/CT angiography | 53.3 (48) |
| Upper endoscopy | 41.6 (37) |
| Colonoscopy | 42.7 (38) |
| Hepatic doppler ultrasound | 38.2 (34) |

### Treatment and Interventions

Medical or therapeutic interventions had been initiated in 27,8% of the population. Iron supplementation was the most frequently administered therapy, with 60% receiving oral iron and 41.1% receiving intravenous iron. Tranexamic acid was used in 24.4% of patients, and propranolol in 11.1%. Among those with hepatic AVMs, 10% had been treated with propranolol.

Regarding interventional procedures, nasal cauterization was performed in 16.7% of patients, and embolization in 6.7%. Additional therapies included folic acid (n = 4) and tamoxifen (n = 1) (Table 7).

**Table 7.** Treatment frequency in the studied population.

| **Treatment** | **Frequency (% , n)** |
| --- | --- |
| Oral iron | 60 (54) |
| IV iron | 41.1 (37) |
| Tranexamic acid | 24.4 (22) |
| Propranolol | 11.1 (10) |
| Cauterization | 16.7 (15) |
| Embolization | 6.7 (6) |
| Folic acid | 4.4 (4) |
| Tamoxifen | 1.1 (1) |

## Discussion

This study estimated a prevalence of 3.83 per 100,000 inhabitants, which is significantly lower than global estimations (19–24), and even more relevant, lower than the only regional estimation performed for Latin America published by the first HHT reference center of excellence in the region located in Argentina (24). Using the same standardized prevalence calculation performed by Serra et al., (using a denominator of 10000 patients and calculating a prevalence of 3.2 in 10000 (24)), our prevalence estimation is an order of magnitude less than theirs (0.383 in 10000 patients) evidencing HHT is also a rare and underestimated health condition in Uruguay. Considering the proximity between Argentina and Uruguay, our shared origin, history, culture and costumes, it would be expected a similar prevalence in this hereditary disease. However, in this case the calculated prevalence is rather low compared to the Argentinian probably in part due to a smaller total population in Uruguay and other relevant factors. Among them, the situation may be attributed to outdated, misleading or incorrect beliefs held by patients (24), not assuming the relevance of the suffered health condition, and by some physicians (24), not recognizing the HHT condition while serving in the first line of consultation, in both cases contributing to a substantial underdiagnosis and underreporting of the cases. It is important to highlight that this is reflected by the reported average diagnostic delay of nearly six years underscoring a critical gap in the disease recognition, timely screening, and access to effective, and sometimes unknown (24), available curative interventions for HHT.

Additionally, another relevant factor could be the lack of accessibility to proper systematic and early genetic testing, something that has been emphasized as a must at least for neonates born to parents with a history of the condition in the HHT international clinical guidelines (1). Currently in Uruguay, few patients have their genetic test performed, most of them abroad, and covered at their own expenses. Notably, in some cases, even the mediation of local HHT-referent physicians have contributed to get access to genetic testing abroad allowing also the chance of even identifying *de novo* cases in patients lacking a HHT family history. This points out the need for implementing a local and broader alternative strategy, maybe following the HHT international guidelines, that help improve national healthcare through the benefit of accessing genetic testing.

Our study aligns with other reports (24,25,26) showing a female predominance (female-to-male ratio 1.78:1). This occurs despite HHT being an autosomal dominant inherited disease with a 50% chance of inheriting the mutated allele of an affected parent, and that this bias has to be attributed to an additional explanation. Interestingly, It was previously suggested that this difference may occur due to females’ higher healthcare utilization (24) influenced by specific reproductive health needs throughout their lives, including menstruation, pregnancy, childbirth, menopause, and other chronic and non-chronic conditions. However, additional research is needed to clarify the source of this bias.

Epistaxis also aligns with the expected values for the disease (90% manifestation in adults (27)), and more than 50% of patients exhibited moderate to severe epistaxis measured by the Epistaxis Severity Score (ESS), highlighting the need for specialized otolaryngologic evaluation and access to effective planned therapies. Although median hemoglobin levels appeared within normal limits, many patients were undergoing iron supplementation or had received transfusions, reflecting chronic iron-deficiency anemia despite treatment.

Gastrointestinal involvement was observed in a significant proportion of patients, which may be possibly associated with the higher regional prevalence of mutations in the gene ACVRL1 (28). The frequency of visceral AVMs differed from global reports (29), likely reflecting the low rate of complete screening in our study population. Only 22% of patients underwent comprehensive imaging in accordance with international screening guidelines (1), limiting accurate detection and classification of systemic involvement. Additionally, the progressive nature of visceral AVMs and their potential complications, and the delayed diagnosis may result in preventable morbidity and mortality.

Our study also considered current treatments being performed in the HHT population. The most common treatments included oral and intravenous iron and antifibrinolytics such as tranexamic acid. Although propranolol, an agent with antiangiogenic properties, is recommended in some cases of hepatic AVMs, only half of the affected patients received it. In the absence of an officially designated national referral center for HHT in Uruguay, it remains challenging to ensure access to recommended treatments. Many of the therapies endorsed by international guidelines, such as bevacizumab (1), are high-cost and not currently approved for use in Uruguay. Their use is further complicated by the rarity of the disease, which limits the availability of traditional forms of clinical evidence typically required to support their indication.

This is the first epidemiologic study of HHT conducted in Uruguay, providing a foundational insight into the national disease burden. This study highlights a clear need to raise awareness about HHT at both clinicians at different levels of response and the broad national healthcare system. Rare diseases such as HHT require management by multidisciplinary teams trained in disease-specific care. Establishing such a team within a formalized national healthcare framework would improve outcomes, reduce preventable complications, optimize resource utilization, and enhance the quality of life for patients and their families. These findings underscore the urgency of national recognition of HHT as a health priority and the creation of a dedicated reference center to ensure equitable access to diagnosis, treatment, and specialized care for all affected individuals in Uruguay.

## Conclusion

HHT is a rare but underdiagnosed disorder in Uruguay, characterized by significant diagnostic delays and limited access to recommended evaluations and treatments. Although a HHT group exists in the country and is working on different aspects in the field, the absence of a formally established national framework contributes to inconsistencies in care and restricts access to comprehensive and multidisciplinary management.

This first epidemiological study of HHT in Uruguay provides essential insights into the clinical and diagnostic landscape of the disease and paves the road for future initiatives aiming to enhance awareness, improving early detection, and ensuring equitable access to evidence-based healthcare for affected individuals.

## Data Availability

All data produced in the present study are available upon reasonable request to the authors.

## List of abbreviations

(HHT): Hereditary Hemorrhagic Telangiectasia
(AVM): arteriovenous malformation
(ESS): Epistaxis Severity Score
(SD): standard deviation
(IQR): interquartile range
(Hb).: hemoglobin

## Declarations

### Ethics approval and consent to participate

Ethical considerations and consent of the participants were included in this study following the approval of the Ethics Committee of Maciel Hospital, Montevideo, Uruguay under the code #13 (2023).

### Consent for publication

Not applicable.

### Availability of data and materials

The dataset used and/or analysed during the current study are available from the corresponding author on reasonable request.

### Authors’ contributions

G.L., B.M., L.M., R.N., H.P., S.R. performed the interviews and analyzed patient’s data. C.C. reviewed the patient’s data and contributed to the table’s organization. Z.C., R.M., S.R. prepared and designed the study, analyzed, interpreted and wrote the manuscript. All authors read and approved the final manuscript.

## Acknowledgements

We want to thank the Uruguayan HTT Patients Association, involved patients and their families for supporting this study. Additionally, we thank the Unidad Académica Médica 1 and Clínica Médica 1 at Hospital Maciel for their support. Z.C. wants to thank Dr. Jorge Facal for his support and advice.

